# Impact of elective inpatient treatments on monthly earnings and employment: a national linked data study in England

**DOI:** 10.1101/2025.11.19.25340566

**Authors:** Hannah Aston, Hannah Bunk, Thomas Henstock, Anushka Madhukar, Svetlana Batrakova, Adam Memon, Vahé Nafilyan, Daniel Ayoubkhani

**Affiliations:** Health and International Directorate, Office for National Statistics, Newport, UK; Strategic Analysis Team, NHS England, London, UK; Strategy Team, NHS England, London, UK

**Keywords:** Treatment effect, Elective surgery, Elective procedures, Labour market, Employment, Unemployment, Pay, Earnings, Income

## Abstract

**Objective:** Evaluate the impact of elective inpatient hospital care on monthly earnings and employment status among working-age adults.

**Design:** Interrupted time series analysis using national, linked administrative datasets.

**Setting:** Hospital inpatient services in England between 1 April 2014 and 31 March 2023.

**Participants:** 5,762,437 individuals who 1) had at least one elective (non-emergency) inpatient admission recorded in NHS Hospital Episode Statistics (HES) starting on or after April 2015 and finishing on or before March 2023; 2) had a treatment specialty code from a specified list; 3) were aged 30 to <59 years at the date of treatment; 4) could be linked to national census and taxation information.

**Main outcome measures:** Monthly employee pay expressed in 2023 prices and paid employee status.

**Results:** We found that elective inpatient treatments were associated with improved long-term economic outcomes relative to a counterfactual scenario of untreated health deterioration across a range of treatment specialties. At 60 months post-treatment, patients treated under the Clinical Haematology Service exhibited the largest treatment effect on earnings: by the end of the five-year outcome period, treated individuals earned £1,805 more per month than the counterfactual. Patients treated under the Clinical Oncology Service showed the largest treatment effect on employment at 60 months, with an employment rate 29.9 percentage points higher than the counterfactual at the end of the outcome period. Across all treatment specialties, the total cumulative effect over the five-year outcome period was £47.2 – 73.7 billion in earnings and 198,918 - 745,262 person-years of employment.

**Conclusions:** Receiving elective inpatient treatment under a range of specialities was found to have a sustained positive impact on both earnings and employment rates across a five-year follow-up period. This suggests that receiving treatment may lead to improved participation in the labour market, contributing to economic growth, and the benefit of this persisting many years after the treatment itself was received.

**Summary box:** *Section 1: What is already known on this topic:*

- Small studies focusing on singular treatments have suggested that there is an improvement in labour market outcomes after treatment for health conditions

*Section 2: What this study adds:*

- Our study suggests that across a range of treatment groups, there is a positive treatment effect on both pay and employment, which would not have been observed had treatment not taken place
- Our study builds on previous research by utilising a linked population-level dataset for England, incorporating electronic health records, sociodemographic information from the national census, and pay data collected for tax purposes, with five years of follow-up across of breadth of treatments

## Introduction

The economic consequence of poor health is a growing concern. Poor health can affect an individual’s capacity to work, thereby influencing employment prospects, income levels and productivity (1). Economic inactivity (defined as not working nor looking for work) due to poor health is rising in the UK. In the first quarter of 2025, 2.8 million people were economically inactive due to long-term sickness in the UK, up from 1.9 million in 2016 (2).

Existing evidence suggests that ill-health is associated with poorer labour market outcomes, including employment and earnings (3) (4). In the UK, people with poor mental health were found to have a 17 percentage point lower employment rate, whilst those who develop a physical impairment had a doubled probability of reduced productivity due to health (5). Between 2023 and 2024, it was estimated that sickness absence accounted for nearly 150 million lost working days, an average of 4.4 lost days per worker (6).

However, there is limited research on how targeted health interventions might improve these outcomes. Demonstrating a clear link between such interventions and economic benefits could support the case for expanding healthcare provision for the working-age population, with potential to enhance labour market participation and contribute to increased economic growth, as well as improving the income and wellbeing of the affected individuals and their households.

A study of over 13,000 men in the US across a six-year period found that treatments to reduce cardiovascular-related mortality led to an increase in earnings and family income (7). In Denmark, a longitudinal study found that receiving bariatric surgery led to a total increase in earnings of 5% and increased labour market participation (8). In the UK, those who received bariatric surgery also saw an average increase in pay of £84 per month in the five years after surgery when compared with the six months prior (9). Another UK study involving a small cohort of individuals who underwent orthopaedic surgery found that nearly all returned to work following surgery, concluding that the treatment was effective in maintaining employment (10). However, the study’s limited sample size and reliance on self-reported outcomes limit the strength of its findings. In contrast, a large-scale study of over 185,000 US adults demonstrated that improved physical function was associated with a higher likelihood of employment (11). Nevertheless, this study is now over a decade old and, like the former, did not isolate the specific impact of treatment itself.

Although there is some evidence linking elective treatment to improved economic outcomes, the research is still relatively limited. Many studies focus on specific conditions or interventions, often with small sample sizes or short follow-up periods. Others rely on self-reported outcomes, which can introduce bias, and few are able to clearly separate the effect of treatment from other factors. There is also a lack of UK-based, population-level evidence covering a broad range of treatments and conditions. As a result, we still know relatively little about how healthcare can support people to stay in or return to work, or what the wider economic benefits might be.

Therefore, this study aimed to evaluate the long-term impact of a range of elective hospital treatments on employment and earnings, using a linked population-level dataset for England, incorporating electronic health records, sociodemographic information from the national census, and pay data collected for tax purposes, with five years of follow-up. To our knowledge, no previous UK-based study has examined the labour market effects across a broad range of treatments for inpatients at the population level. Unlike earlier research, our approach benefits from timely, longitudinal data and allows for direct measurement of labour market outcomes through tax records, rather than relying on retrospective questionnaires or other proxies.

## Methods

### Study design and data sources

We conducted an observational, retrospective, longitudinal study of individuals who received inpatient elective treatment between 1 April 2015 and 31 March 2023 (the latest available hospital records to us at the time of analysis), under one or more of 32 treatment specialities from a pre-specified list of the most common treatments in inpatient activity (small counts precluded analysis of other specialties). Collectively, these treatment specialties accounted for approximately 96% of inpatient activity (excluding likely childbirth-related activity) among working-age patients in hospital records in 2023/24.

We created a person-level dataset combining Hospital Episode Statistics (HES) Admitted Patient Care (APC) (12) records with: sociodemographic characteristics from Census 2011 (13); Office for National Statistics (ONS) death registration data (14); and Pay As You Earn (PAYE) Real Time Information (RTI) records from His Majesty’s Revenue and Customs (HMRC). These are records of gross earnings paid to employees collected for tax purposes for the UK government. These payment records were calendarised to monthly longitudinal observations on employee status and pay (15).

The analysis made use of the 2011 Census rather than the 2021 Census, as this was the most recently available census data at the start of the study period (1 April 2015). Use of the 2021 Census could have introduced survivorship bias into the analysis.

The PAYE dataset was linked to the 2011 Census through the ONS Demographic Index (16), an anonymised, person-specific identifier compiled by ONS from various national administrative data sources. The HES and death registration datasets were joined to the 2011 Census by the NHS number, obtained via linkage of the 2011 Census and NHS Patient Registers 2011-2013 (17). All datasets were then de-identified, prior to being integrated and analysed.

### Study population

For each treatment specialty, the study population included people who were enumerated at the 2011 Census and were usual residents of England, could be linked to NHS and HMRC records, and had a HES record that met all of the following criteria: an admission date between 1 April 2015 (one year after the start of the HMRC PAYE dataset) and 31 March 2023, to align with the latest available PAYE data at the time of analysis; a treatment specialty code corresponding to the specialty of interest; an elective (non-emergency) admission; a waiting time of two years or less between the decision to admit and admission date, with longer waits considered likely a result of data error; and an age between 25 and 64 years at the time of admission.

The dataset was further filtered to only include monthly data when patients were aged between 30 and 59 to compute the age-standardised values (see ‘statistical analysis’ below). This is to ensure that people in all age groups that were present in the study population in the month of surgery could contribute to the age-standardised estimates in all months before and after surgery, before “ageing out” of the age range covered for follow-up. For example, if people aged 60 to 64 years in the month of surgery were retained in the study population, they would have “aged out” of follow-up by 60 months post-surgery, hence they would not contribute to the age standardisation calculation in these months, and the age-standardised estimates would appear artificially low. The same logic applies to people aged 25 to <30 years in the month of surgery.

Within each treatment specialty, patients who had a HES record from 1 April 2009 (the earliest available HES data) to 31 March 2015 under the corresponding treatment specialty were excluded from the study population. This increased the likelihood that patients included in the study population were being treated for a health condition for the first time within the study period. In instances where a patient had more than one qualifying record during the study period, the record with the earliest admission date was kept. The date of this admission was designated the ‘index date’ for follow-up.

To compare labour market trajectories of patients in each of the study cohorts to that of the general population, a 1% sample was randomly selected from the enumerated 2011 Census population who were usual residents of England, could be linked to NHS and HMRC records, and were alive on 1 April 2015 (i.e. no linked death registration before this date). Index dates were randomly assigned following the distribution of admission dates in the study population across all treatment specialties combined.

### Study period and follow-up

For each of the study populations, plus the Census-based general population cohort, an analytical dataset was curated with one row per person per month. The calendar month containing the index date was assigned ‘Month 0’. Patients were followed for up to 60 calendar months (i.e. five years) before and after Month 0. Follow-up was censored when patients “aged out” of the study population (in either direction, < 30 years or ≥ 59 years); after they had died; before 1 April 2014 (start of the PAYE dataset); and after 31 March 2023 (end of the PAYE dataset).

### Exposure

The exposure was time before or after the first inpatient procedure.

### Outcomes

The labour market outcomes of interest included gross pay and paid employee status. Gross pay was derived from the HMRC PAYE dataset, calendarised to monthly values, winsorised at the 99.9th percentile to reduce the influence of extreme values (which could be due to measurement errors) and deflated to 2023 prices using the Consumer Price Index including owner occupiers’ housing costs (CPIH). Individuals without a record in the PAYE dataset for a given month were imputed to have earned £0 in that month. Paid employee status was defined as a binary indicator, where an individual was considered a paid employee if their gross monthly pay (after imputation) exceeded £0.

Outcomes were derived over the period 1 April 2014 to 31 March 2023. The outcome period (defined by the availability of PAYE data) starting one year earlier than the study inclusion period means that all included patients had at least one year of pre-treatment follow-up (unless they “aged out” of the study population during this period).

### Statistical analysis

This study employed an interrupted time series approach. For each treatment specialty, we estimated mean monthly earnings and employment rates by month, both prior to and following treatment, standardising for age using the age distribution at the time of treatment. This age standardisation is essential to account for age as a key time-varying confounder, given that labour market outcomes evolve with age due to factors such as retirement, transitions to part-time work, or earnings progression. This was done by standardising monthly values of mean pay and probability of employment, such that the age distribution (measured in half-year intervals) in each month was held fixed at that in Month 0.

To derive estimates of treatment and exposure effects, linear regression models were fitted to age-standardised values of pay/employment over a span of the pre-treatment period. As the study period for HES inclusion began on March 2015 and the PAYE data went back as far as March 2014, we only used the one-year period before treatment for model fitting, ensuring all the study population had the opportunity to contribute to the estimated trajectory. The one-year period before treatment was conceptually divided into two epochs: ‘pre-disease’ (i.e. not yet ill) and ‘pre-treatment’ (i.e. ill but not yet treated), the transition to the ‘pre-treatment’ phase was our breakpoint. The location of the breakpoint was automatically identified using R’s ‘segmented’ package (18). This is done by fitting piecewise linear regression models using an iterative algorithm that estimates the location of change-points (breakpoints) where the relationship between the predictor and response variable shifts (19).

These models were then used to project pay/employment outcomes in the absence of treatment, by extrapolating the fitted trajectory from the ‘pre-treatment’ epoch. For every month up to five years after the treatment, the effect of the treatment was estimated as the difference between the observed and predicted age-standardised monthly earnings and employment rate.

To capture uncertainty in both age-standardised estimates and counterfactual pay/employment values, confidence intervals (CIs) were generated via simulation, as bootstrapping was computationally infeasible. For each of the study populations and 121 time periods (Month −60 to Month 60), 10,000 values of pay among paid employees were simulated from a log-normal distribution, and 10,000 probabilities of being a paid employee were simulated from a binomial distribution, both based on observed data. We then multiplied the simulated values for pay among paid employees by the simulated probabilities of being a paid employee to derive simulated values of pay among all working-age people. Simulated values were age-standardised, standard errors were calculated as the standard deviation across simulations and 95% CIs were then constructed as: estimate ± 1.96 × SE.

As the 32 study cohorts are not mutually exclusive, some of the estimated cumulative treatment effects in pay/employment for a given treatment specialty may be attributable to patients being treated in other specialities. Thus, there will be some degree of double-counting if estimates of the cumulative effects are simply summed across treatment specialties, giving an upper bound. To provide a more conservative estimate, cohort sizes were re-calculated under the assumption that patients belonging to multiple study cohorts accrued for just one specialty in which they were treated: the one with the smallest per-person effect size. The re-calculated cohort sizes were then multiplied by the per-person effect sizes and summed across the 32 treatment specialties, giving a lower bound on the estimated cumulative effect.

Further detail on the statistical methods can be found in the supplementary appendix.

### Sensitivity analysis

In addition to performing a breakpoint analysis with linear regression models, we also explored second- and third-order polynomial regression models fitted to the entire one-year pre-treatment span, thus allowing for a gradual rather than an abrupt change in the trajectory.

Although these models sometimes provided a better fit to the training data than the linear breakpoint models, they often resulted in “explosive behaviour” in their out-of-sample projections, with estimated values of mean pay and probability of paid employment rapidly tending to zero after Month −1 (see the supplementary material). These models were therefore not considered further.

### Statistical software

All analyses were carried out in R version 3.5.1. Dataset linkage and cleaning was carried out using Spark version 2.4.0.

### Patient and Public Involvement

Due to the wide range of treatment specialties covered, it was not possible to involve patients and the public in this research.

## Results

### Characteristics

Between 1 April 2015 and 31 March 2023, 4,867,218 study-eligible individuals were admitted to hospital for an elective inpatient treatment under one or more of the 32 specialities covered in our analysis. The sample flow is provided in Supplementary Figure 1. The results presented in this section cover the top 20 most common treatment specialties in HES in 2023/24. Data for all 32 treatment specialties covered in the analysis are available in the supplementary materials and on the ONS website (20).

The mean age of the study population was 47.7 years, 54.1% of individuals were female, and 81.9% were White British.

Across the study period, 4,508,358 (74.1%) of inpatients in our study population received treatment under one speciality. A total of 1,185,421 (19.5%) of patients received treatment under two specialities and 299,485 (4.9%) received treatment under three or more specialties.

The treatment specialty with the highest number of admissions was the Gastroenterology Service, with 1,447,590 admissions recorded over the study period. In contrast, the specialty with the lowest number of admissions was the Renal Medicine Service, which accounted for 18,934 admissions. See the supplementary materials for the most common diagnosis and procedure codes within each of the treatment specialities covered.

### Treatment effects on earnings and employment at the end of follow-up

Earnings and employment outcomes varied across inpatient treatment specialties. Under the counterfactual model, average earnings generally declined over time, while the observed earnings trajectories following treatment either stabilised or increased (Figure 1). Similarly, the employment rate would generally have steadily decreased without treatment, whereas observed trajectories following treatment were generally higher (Figure 2). In all cases, differences are reported relative to a counterfactual model in which treatment did not occur and health deteriorated over time.

**Figure 1.**
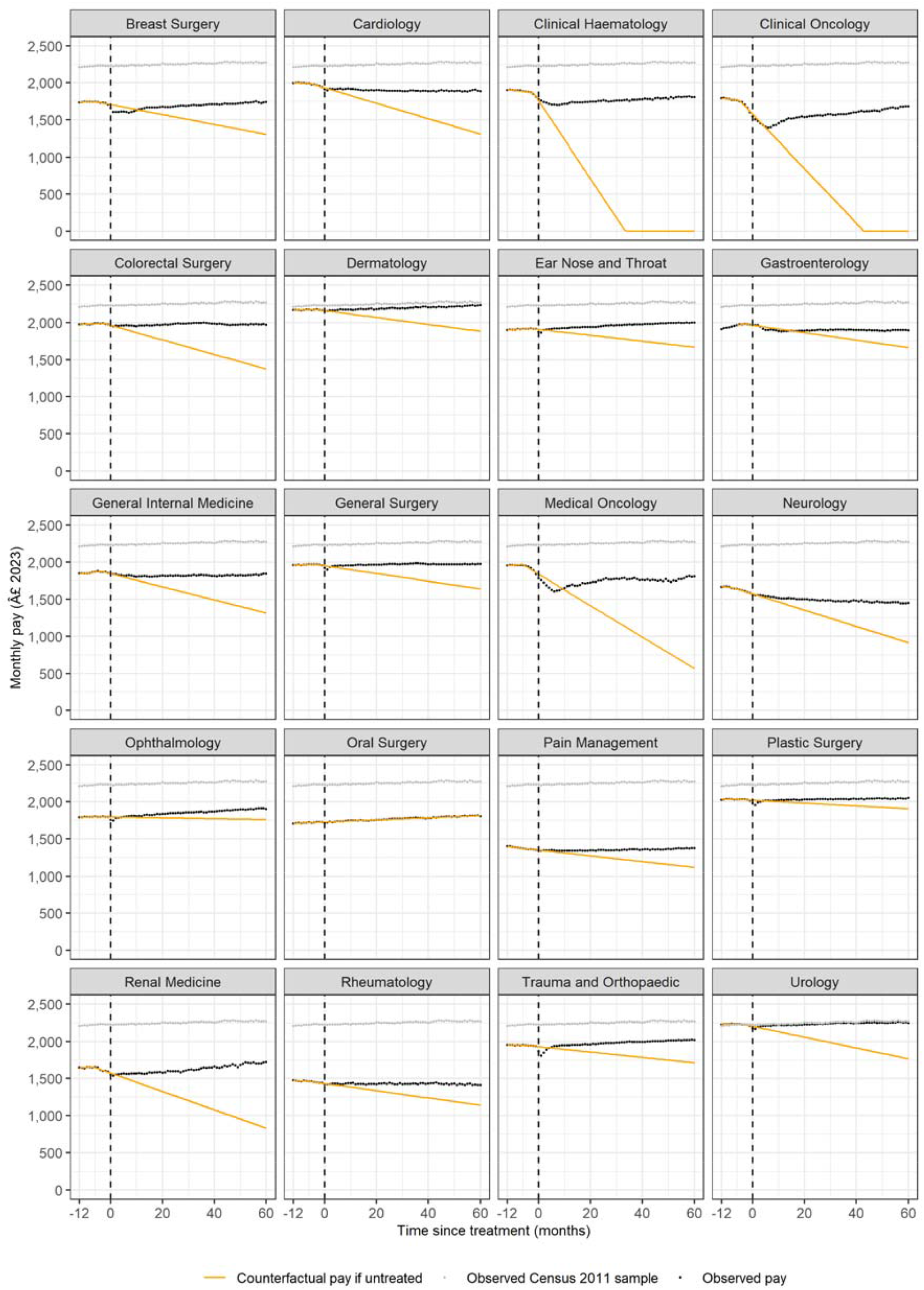
Age-standardised trajectories of pay for the top 20 most common treatment specialities

**Figure 2.**
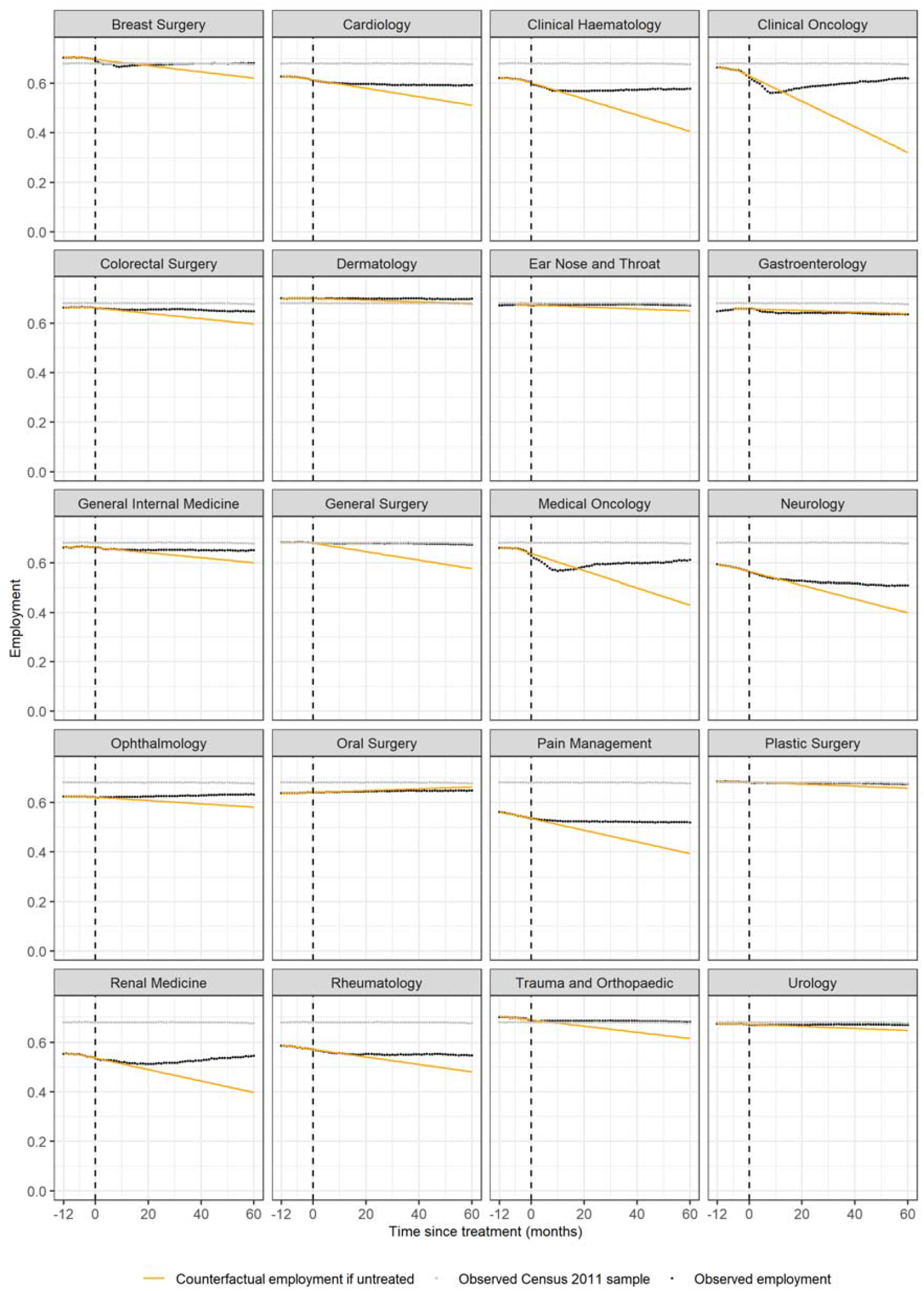
Age-standardised trajectories of employment for the top 20 most common treatment specialities

At 60 months post-treatment, patients treated under the Clinical Haematology Service had the largest estimated treatment effects: on average, treated individuals earned £1,805 more per month than the counterfactual (95% CI: £1,754 to £1,857), and their employment rate was 17.1 percentage points higher (95% CI: 16.0 to 18.2). Patients treated under the Clinical Oncology Service and Medical Oncology Service also experienced substantial gains versus the counterfactual, with monthly earnings increasing by £1,680 (95% CI: £1,631 to £1,730) and £1,239 (95% CI: 1,180 to £1,299), respectively; and employment rates increased by 29.9 (95% CI: 28.7 to 31.1) and 18.2 (95% CI: 16.9 to 19.5) percentage points, respectively.

Patients treated under the Renal Medicine Service, the Colorectal Surgery Service, the Cardiology Service, the Neurology Service and the General Internal Medicine Service all saw increases in average earnings increases of over £500 a month at 60 months of follow-up: £896 (95% CI: £809 to £983), £594 (95% CI: £575 to £614), £579 (95% CI: £553 to £606), £535 (95% CI: £495 to £574), and £524 (95% CI: £500 to £547) respectively. Of these five treatment groups, those treated under the Renal Medicine Service saw the biggest increase in their employment rate at 60 months post-treatment versus the counterfactual of no treatment at 14.8 percentage points (95% CI: 12.9 to 16.6)

Specialities with estimated treatment effects on average monthly earnings under £500 included the Urology Service, the Breast Surgery Service, the Dermatology Service, the General Surgery Service, the Ear Nose and Throat Service, and the Trauma and Orthopaedic Service.

All other treatment specialties within the top 20 most common (the Rheumatology Service, the Pain Management Service, the Gastroenterology Service, the Plastic Surgery Service and the Opthamology Service) had an effect on average monthly earnings at 60 months lower than £300. Despite a relatively low effect on earnings, those treated under the Pain Management Service saw an increased employment rate of 12.6 percentage points (95% CI: 12.0 to 13.2) at 60 months post-treatment versus the counterfactual of no treatment.

### Cumulative effect on earnings and employment after five years of follow-up

Across the five-year follow up period, the largest cumulative (across all patients and time periods) treatment effects on earnings were estimated for General Surgery (£8.8 billion), the Gastroenterology Service (£7.5 billion), the Trauma and Orthopaedic Service (£6.9 billion), the Colorectal Surgery Service (£6.2 billion) and the Urology Service (£5.5 billion). For employment, the largest cumulative treatment effects were estimated for the General Surgery Service (215,030 person-years), the Trauma and Orthopaedic Service (123,635 person-years), the Pain Management Service (42,802 person-years), the Colorectal Surgery Service (41,437 person-years) and the Clinical Oncology Service (36,307 person-years). See Table 2 for cumulative five-year treatment effects for the top 20 most common specialties considered in the analysis.

**Table 1.**
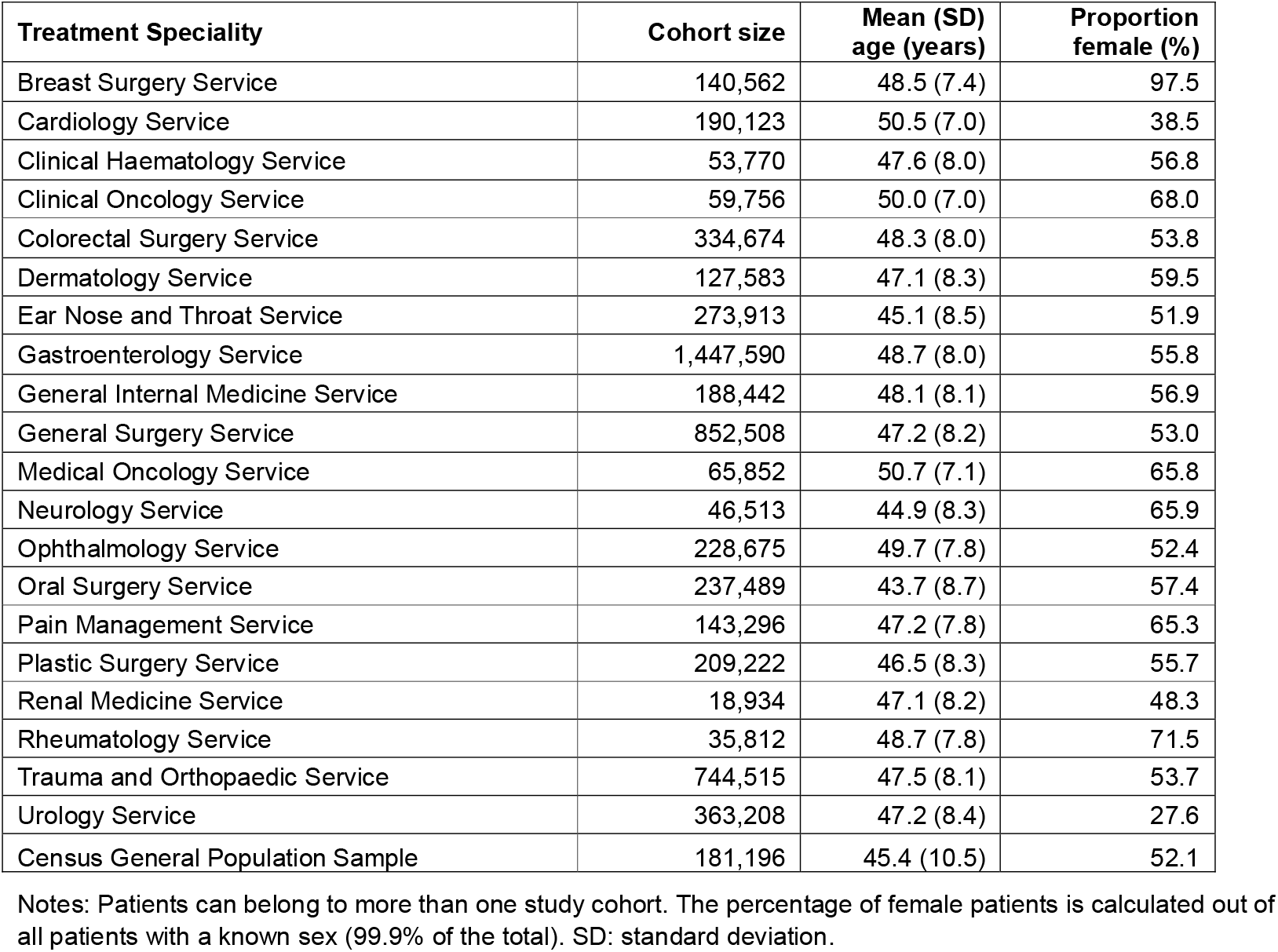
Summary statistics for the top 20 most common treatment specialties.

| Treatment Speciality | Cohort size | Mean (SD)<br>age (years) | Proportion<br>female (%) |
| --- | --- | --- | --- |
| Breast Surgery Service | 140,562 | 48.5 (7.4) | 97.5 |
| Cardiology Service | 190,123 | 50.5 (7.0) | 38.5 |
| Clinical Haematology Service | 53,770 | 47.6 (8.0) | 56.8 |
| Clinical Oncology Service | 59,756 | 50.0 (7.0) | 68.0 |
| Colorectal Surgery Service | 334,674 | 48.3 (8.0) | 53.8 |
| Dermatology Service | 127,583 | 47.1 (8.3) | 59.5 |
| Ear Nose and Throat Service | 273,913 | 45.1 (8.5) | 51.9 |
| Gastroenterology Service | 1,447,590 | 48.7 (8.0) | 55.8 |
| General Internal Medicine Service | 188,442 | 48.1 (8.1) | 56.9 |
| General Surgery Service | 852,508 | 47.2 (8.2) | 53.0 |
| Medical Oncology Service | 65,852 | 50.7 (7.1) | 65.8 |
| Neurology Service | 46,513 | 44.9 (8.3) | 65.9 |
| Ophthalmology Service | 228,675 | 49.7 (7.8) | 52.4 |
| Oral Surgery Service | 237,489 | 43.7 (8.7) | 57.4 |
| Pain Management Service | 143,296 | 47.2 (7.8) | 65.3 |
| Plastic Surgery Service | 209,222 | 46.5 (8.3) | 55.7 |
| Renal Medicine Service | 18,934 | 47.1 (8.2) | 48.3 |
| Rheumatology Service | 35,812 | 48.7 (7.8) | 71.5 |
| Trauma and Orthopaedic Service | 744,515 | 47.5 (8.1) | 53.7 |
| Urology Service | 363,208 | 47.2 (8.4) | 27.6 |
| Census General Population Sample | 181,196 | 45.4 (10.5) | 52.1 |

Notes: Patients can belong to more than one study cohort. The percentage of female patients is calculated out of all patients with a known sex (99.9% of the total). SD: standard deviation.

**Table 2.**
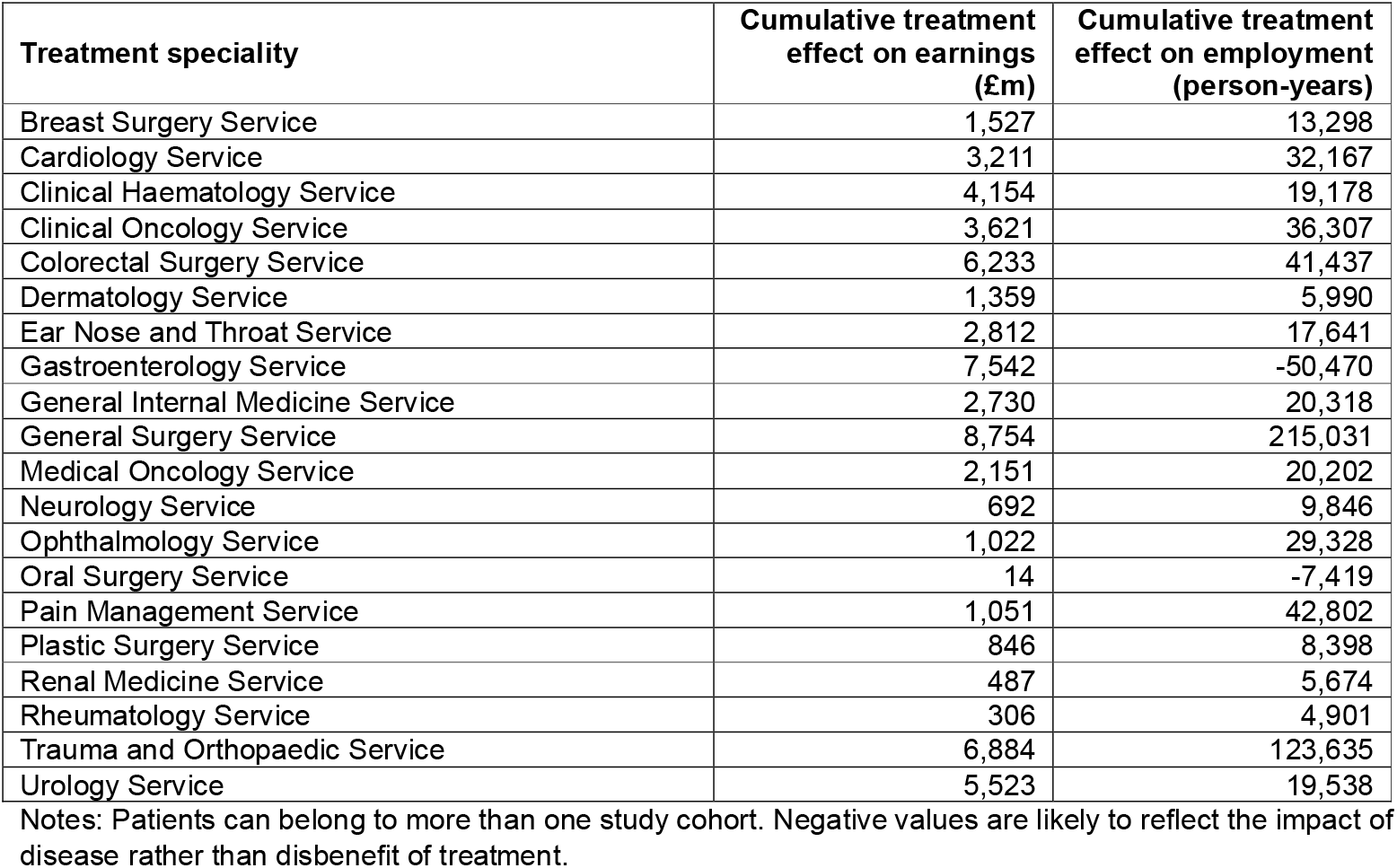
Cumulative treatment effects across the five-year follow-up period.

Across all 32 analysed treatment specialties and ignoring any double-counting of patients between specialties, the cumulative five-year treatment effect summed to £73.7 billion for earnings and 745,262 person-years for employment. These totals represent upper bounds on the estimated total treatment effect.

After adjusting the cohort sizes to account for some patients being treated under multiple specialties and multiplying by the estimated per-person effect sizes, the total cumulative five-year treatment effect across all 32 specialties was conservatively estimated at £47.2 billion for earnings and 198,918 person-years for employment. These totals represent lower bounds on the estimated total treatment effect.

## Discussion

### Principal findings

We found that elective inpatient treatments were associated with improved long-term economic outcomes relative to a counterfactual scenario of untreated health deterioration across a range of treatment specialties.

At 60 months post-treatment, patients treated under the Clinical Haematology Service exhibited the largest treatment effect on earnings: treated individuals earned £1,805 more than the counterfactual, and their employment rate was 17.1 percentage points higher. Patients treated under the Clinical Oncology Service experienced the largest treatment effect on employment at 60 months, with an employment rate 29.9 percentage points higher and earnings £1,680 higher.

Across the full five-year follow-up period, patients treated under the General Surgery Service experienced the largest cumulative treatment effect for both earnings and employment, totalling £8.8 billion and 215,031 person-years, respectively, over the period. The total effect across all treatment specialties was estimated at between £47.2 billion and £73.7 billion for earnings, and between 198,918 and 745,262 person-years for employment.

### Research in context

The positive effect of treatment on earnings observed across treatment groups suggests that improvements in health following elective care may enhance individuals’ ability to engage in the labour market. One plausible mechanism is that treatment restores physical and/or mental health, enabling individuals to return to work or preventing them from dropping out of work in the first place. Another possible mechanism is that the treatment prevents people from reducing their hours or moving to less demanding and lower paid jobs. Both mechanisms align with existing evidence showing that better health is associated with higher labour market participation and earnings.

A substantial body of research has established a strong link between health and quality of life. Over the past five decades, the field of health-related quality of life (HRQoL) has grown significantly, with the number of published studies increasing from virtually none to over 17,000, according to a recent review (21). This expansion reflects a growing recognition of the multifaceted ways in which health affects individuals’ lives.

The consequences of poor health extend beyond personal well-being, influencing labour market outcomes and financial stability. Financial burden is increasingly recognised as a critical aspect of cancer care and patient outcomes (22). For example, evidence from Washington State shows that cancer patients are 2.7 times more likely to declare bankruptcy than individuals without cancer (23). Moreover, financial strain is significantly associated with lower baseline quality of life (24). Poor health can affect an individual’s capacity to work (1) and research consistently shows that individuals in better health are more likely to work and remain employed. In Iran, for instance, nurses with higher HRQoL reported increased labour supply (25). International evidence similarly supports the idea that improved health enhances labour supply and productivity (4), while deteriorating health negatively affects these outcomes. In the UK, women aged 25 to 54 in England who were diagnosed with endometriosis experienced a statistically significant average decrease in monthly earnings (26).

Healthcare and treatments have also been shown to positively influence labour market outcomes. In Indonesia, men treated for iron deficiency experienced improvements in both physical health and economic success (27). In Finland, rehabilitative psychotherapy was significantly associated with better labour market outcomes (28). In England, individuals who underwent bariatric surgery saw an average monthly pay increase of £84 in the five years following surgery, compared to the six months prior (9). Treatment through NHS Talking Therapies was also linked to sustained improvements in labour market outcomes (29), and shorter waiting times for psychological therapies were associated with reduced productivity loss (30).

Thus, our research broadly aligns with existing evidence, demonstrating a sustained positive impact of treatment for health problems on labour market outcomes. Nonetheless, much of the current literature is constrained—often focusing on single interventions, relying on small sample sizes, short follow-up periods, self-reported data, or treatments conducted outside the UK and the NHS context. Evidence on the economic and labour market impact of elective inpatient treatments remains limited and mixed. Some studies report negative effects, such as increased odds of not returning to work after cancer treatment (31), while others find positive effects restricted to specific treatment groups or based on data from outside the UK (9,25,26,29,30).

We contribute to the evidence base in several important ways. First, to our knowledge, this is one of the first studies to examine a wide range of treatments rather than focusing on a single intervention. This enables us to capture variation in treatment effects across different areas of elective care. Second, we use administrative data covering nearly the entire population of England, which reduces bias, increases sample size, and allows for a five-year follow-up period—providing a longitudinal perspective on treatment impacts.

### Strengths and limitations

To our knowledge, this is the largest study internationally to quantify the labour market effects of receiving elective inpatient treatment. This investigation was made possible by a new national linked dataset comprising electronic health records, demographic information from the decennial census, and national taxation records, which is the first linked dataset containing labour market and health data with near-complete coverage for individuals in England. A key strength of this dataset is the incorporation of administrative data, which allows for a large and representative study population including information on their real-world health care utilisation and labour market outcomes, enhancing the validity of our findings.

However, despite these strengths, there are important limitations to consider that contribute to the likely under-estimation of our treatment effects. The study population was restricted to individuals aged 30-59 years, which means that the aggregate treatment effects may be underestimated compared to what might be observed across the full working-age population, which is generally considered to be 16-64 years in the UK (32). This age restriction also limits the generalisability of the findings to younger and older individuals, who may experience different labour market trajectories.

In addition, due to our data source for earnings being HMRC PAYE, people who were not payrolled employees (e.g. the self-employed) were included in the study population but they were treated as not working and earning £0 in the outcome measures. Our estimated treatment effects therefore only capture the contribution of employees. For context, approximately 13% of employed people in the UK were self-employed at the end of 2024 (33). We also did not have information on hours worked or benefits received, such as sick leave and disability benefit, which could be impacted by receiving treatment.

Furthermore, the estimated treatment effects capture economic benefits for individuals (i.e. higher earnings) but no other potential impacts to the broader economy, such as increased output/productivity within firms, reduced welfare benefit payments, and reduced healthcare utilisation costs.

Another limitation of this study is the use of a broad categorisation for the treatment received, defined according to the specialty of the clinician performing the procedure. This allowed us to cover a wide range of treatment groups. However, this limited our ability to account for potentially important nuances such as the specific medical condition being treated, the severity of the condition, or the intensity and duration of the treatment provided. Patients grouped under the same treatment specialty could have very different conditions and needs, which may affect outcomes in ways our analysis does not capture.

A final limitation concerns the counterfactual model we used to estimate what might have happened if treatment had not been given. Although we chose the most suitable type of model and trained it on an appropriate period, by its very nature a modelled counterfactual outcome cannot be empirically tested and validated against observed data. To reduce this uncertainty, we tested three different modelling approaches (segmented linear regression and second- and third-order polynomial fits) and selected the one that provided the most plausible post-treatment pay and employment trajectories. Nonetheless, it is important to acknowledge that any counterfactual estimate is inherently uncertain.

## Conclusions

Receiving elective inpatient treatment under a range of treatment specialities was found to have a sustained positive impact on both earnings and employment across a five-year follow-up period. This suggests that receiving treatment may lead to improved participation in the labour market, and the benefit of this persisting many years after the treatment itself was received Given that waiting times for elective care in England have grown over the past decade (34), and there remains a backlog of patients waiting for treatment following the COVID-19 pandemic (35), increasing provision of elective procedures therefore has the potential to reduce economic inactivity and increase economic growth – a key policy aim of the current UK government (36). Using the results of this study, NHS England has estimated that if waiting times for elective procedures returned to their statutory target (92% of patients treated within 18 weeks of referral), total earnings would increase cumulatively by £2.7 billion and employment by 63,000 person-years between now and the end of 2030/31 (37).

Future work should aim to further disaggregate the treatment groups, such as by specific conditions or procedures, or according to socio-demographic characteristics.

## Ethics statement

This study was ethically self-assessed against the ethical principles of the National Statistician’s Data Ethics Advisory Committee (NSDEC) using NSDEC’s ethics self-assessment tool. We subsequently engaged with the UK Statistics Authority Data Ethics team, who were satisfied that no further ethical approval was required.

## Data availability statement

The linked data sources used in this study are not publicly available and are subject to controlled access due to their sensitive nature. Researchers can apply to access Hospital Episode Statistics data via NHS England’s Data Access Request Service (DARS): https://digital.nhs.uk/services/data-access-request-service-dars

## Acknowledgements

The authors would like to thank David Goll at Number 10 Data Science, and Chris Worsfold and Henry Foster at the Department of Health and Social Care, for their feedback on the study design. We are also grateful to the following national clinical directors and specialty advisors at NHS England for their valuable insights when interpreting the study results: Dawn Adamson, Mark Cheetham, Jeremy Davis, Jonathan Fuld, Harriet Gordon, Martin Heaton, Tracey Irvine, Lesley Kay, Sue Mann, John McGrath, Adam Millican-Slater, Niranjanan Nirmalananthan, Nick Phillips, Simon Ray, Colin Rees, Alexandra Severns, Doug West.

## Footnotes

DA and VN designed the study. HA and HB performed the data analysis. All authors contributed to interpretation of the study results. HA wrote the first draft of the manuscript. All authors reviewed and revised the manuscript and approved the final version.

