## Supplementary Materials for "Impact of elective inpatient treatments on monthly earnings and employment: a national linked data study in England"

**Supplementary Figure 1. Sample flow diagram**

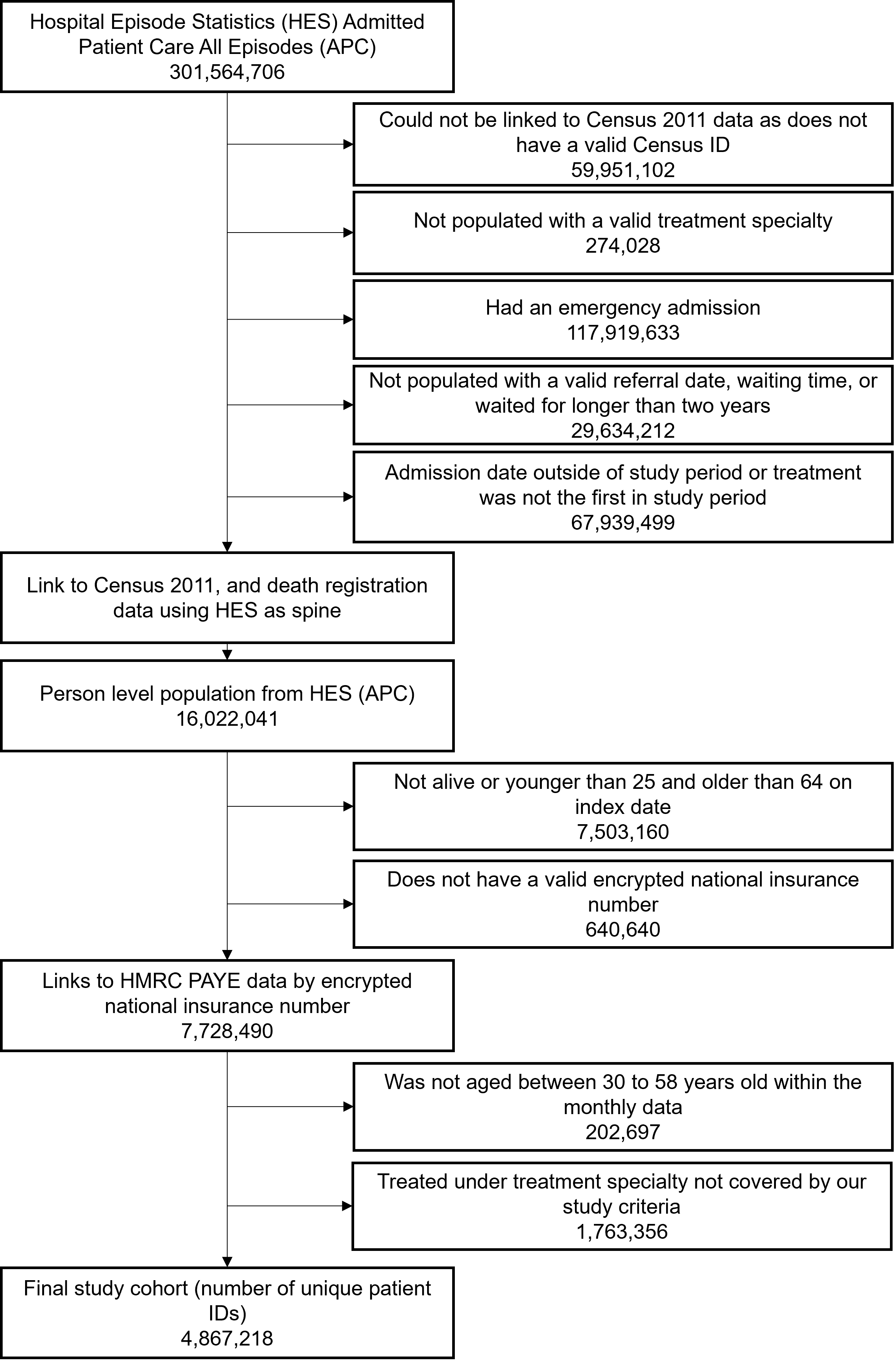

**Supplementary Figure 2. Age-standardised trajectories of pay for the top 20 most common treatment specialities with second order polynomial counterfactual model**

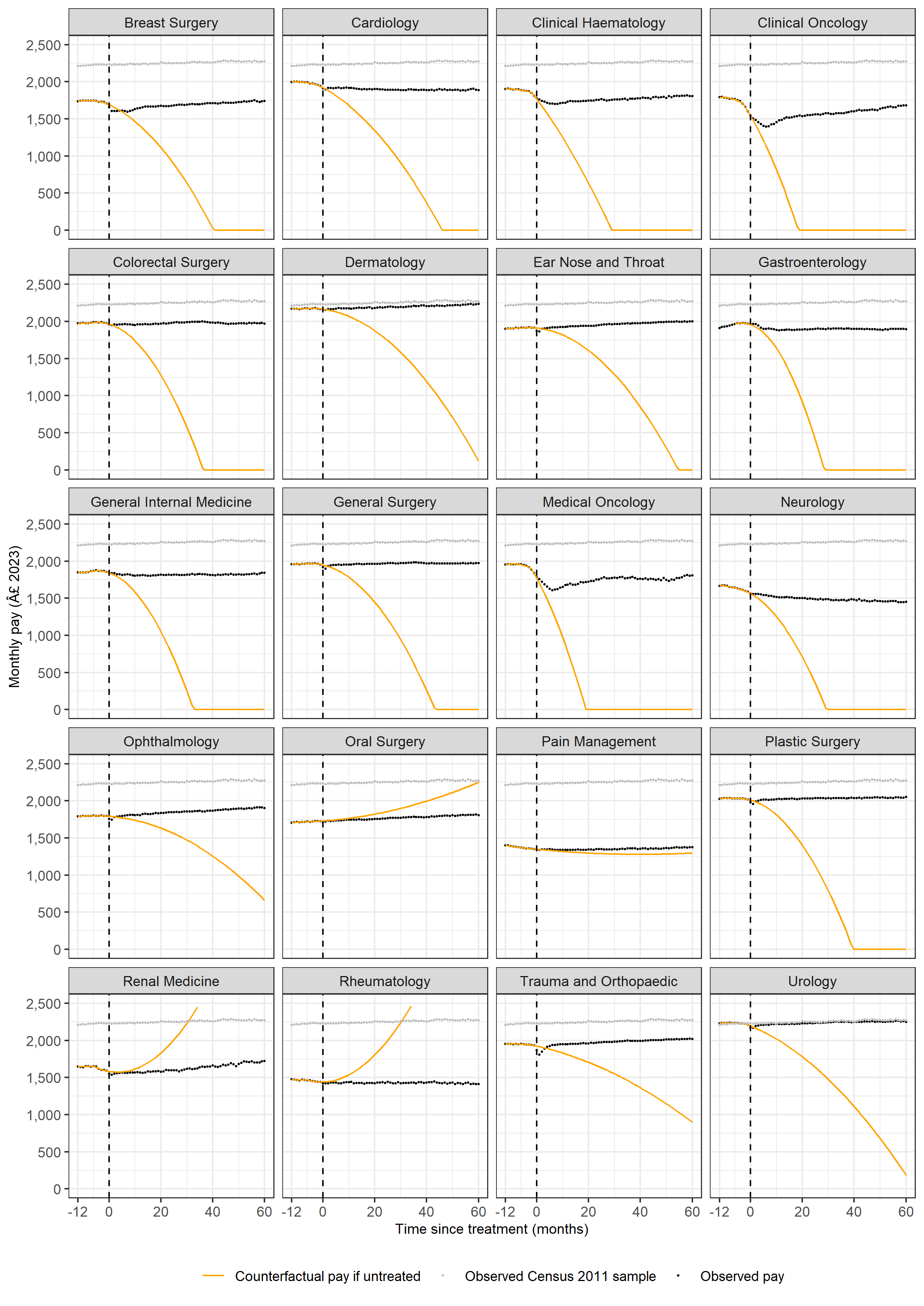

**Supplementary Figure 3. Age-standardised trajectories of employment for the top 20 most common treatment specialities with second order polynomial counterfactual model**

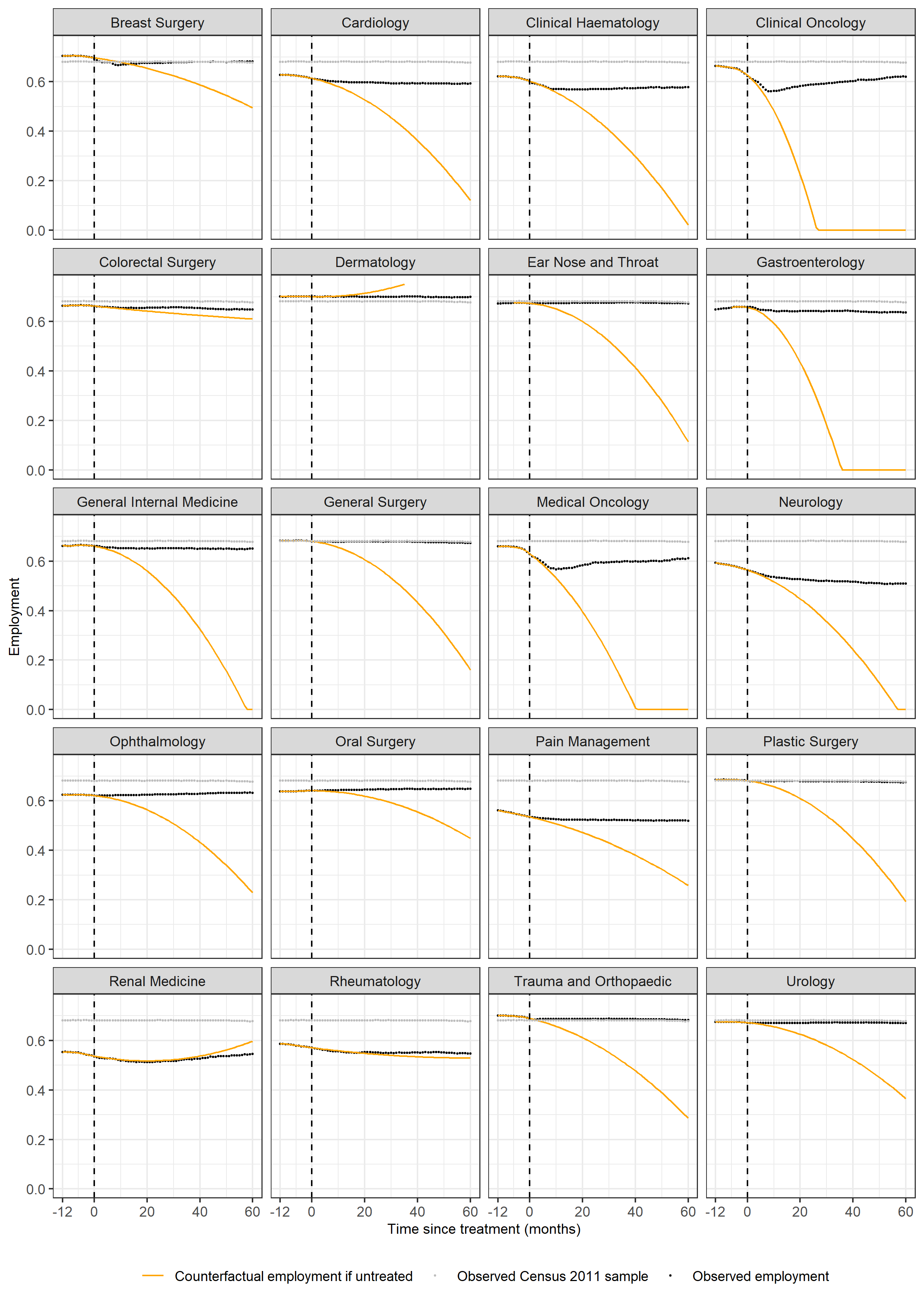

**Supplementary Figure 4. Age-standardised trajectories of pay for the top 20 most common treatment specialities with third order polynomial counterfactual model**

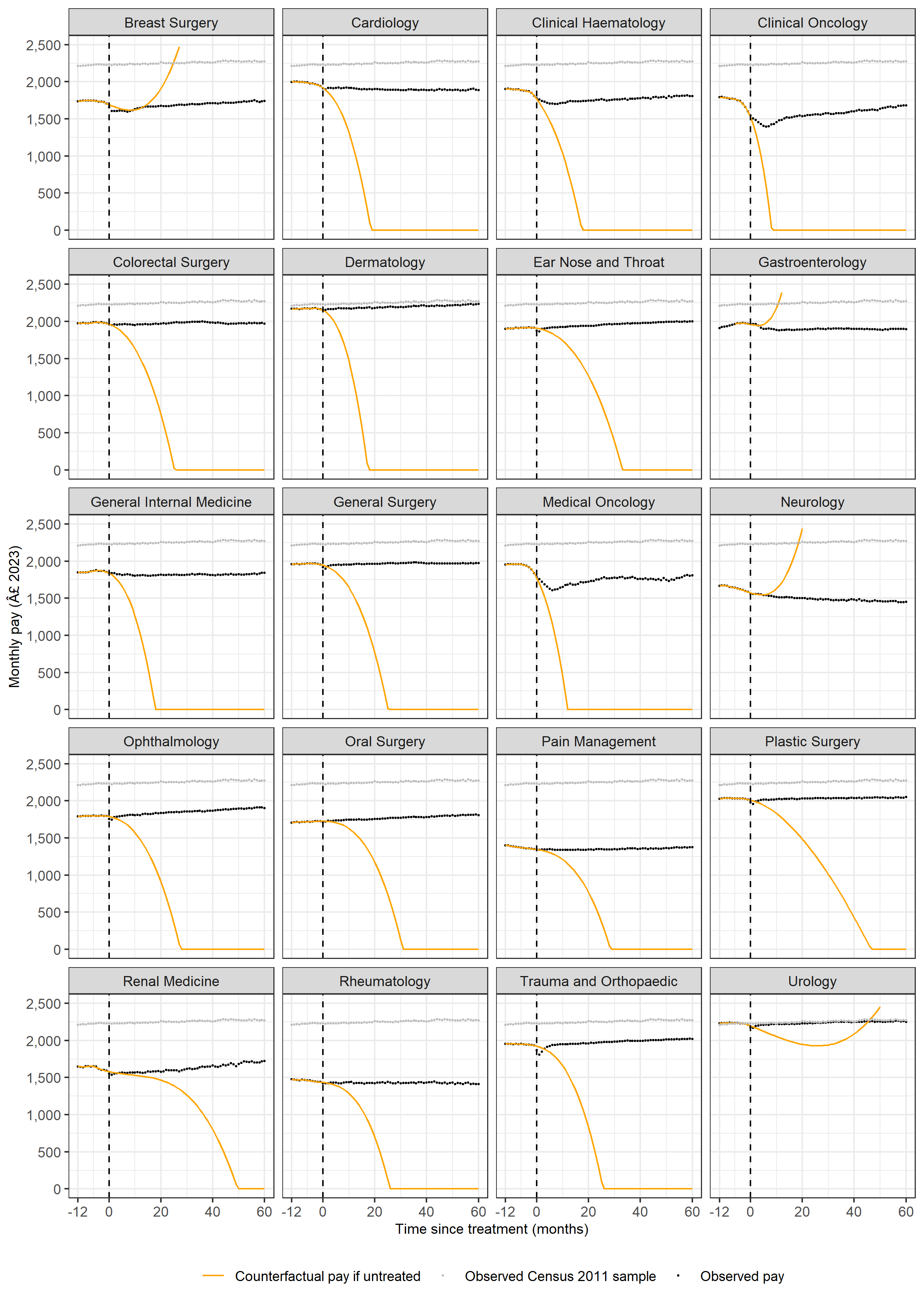

**Supplementary Figure 5. Age-standardised trajectories of employment for the top 20 most common treatment specialities with third order polynomial counterfactual model**

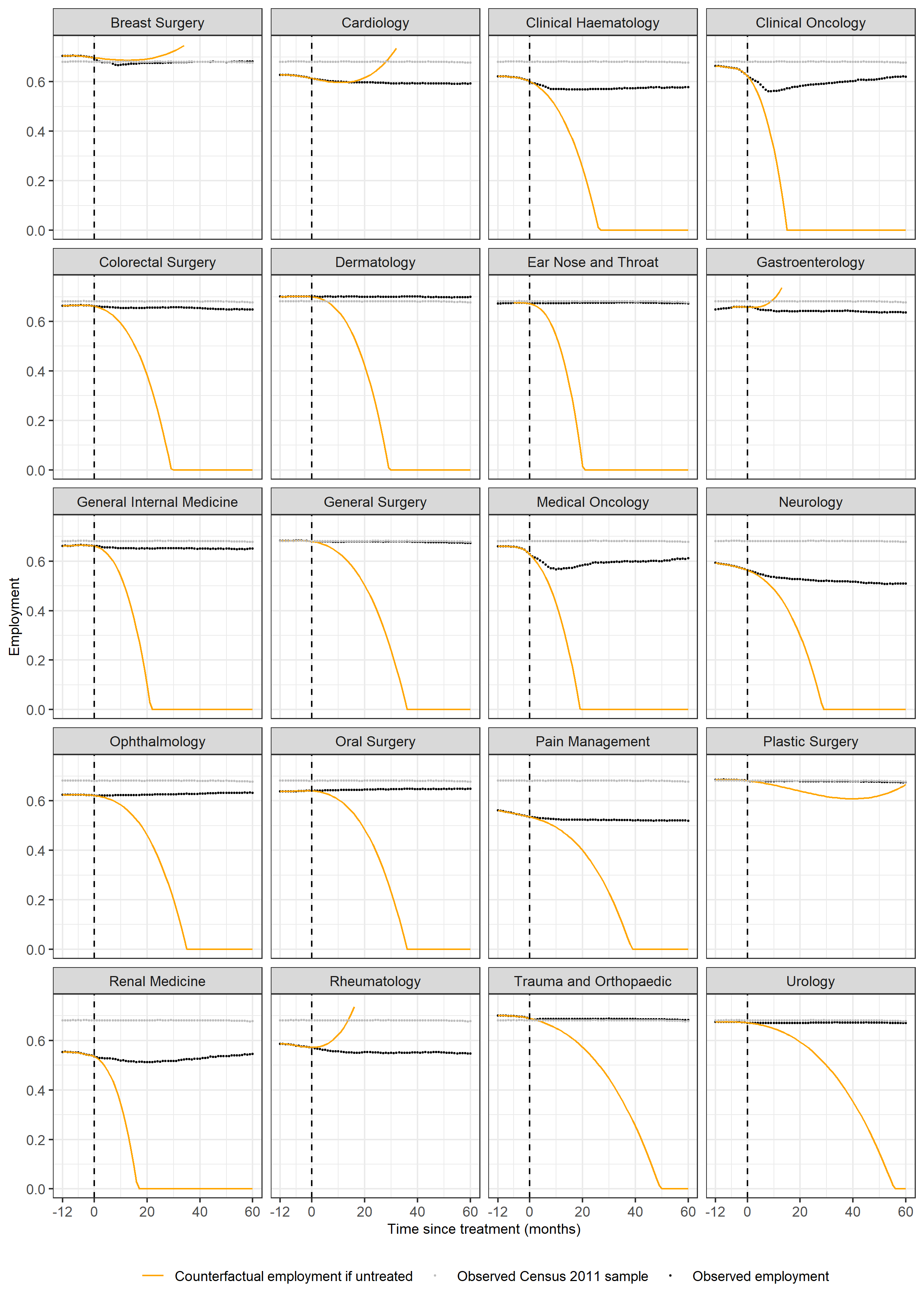

**Supplementary Table 1. Per person treatment effect at 60 months for earnings and employment across all 32 treatment specialities**

| **Treatment specialty description** | **Difference in pay (£)** | **Difference in pay lower 95% confidence limit (£)** | **Difference in pay upper 95% confidence limit (£)** | **Difference in percentage in paid employment (percentage points)** | **Difference in percentage in paid employment lower 95% confidence limit (percentage points)** | **Difference in percentage in paid employment upper 95% confidence limit (percentage points)** |
| --- | --- | --- | --- | --- | --- | --- |
| General Surgery Service | 331 | 322 | 341 | 9.7 | 9.5 | 9.9 |
| Urology Service | 484 | 467 | 500 | 2.2 | 1.9 | 2.5 |
| Breast Surgery Service | 436 | 413 | 459 | 6.0 | 5.5 | 6.6 |
| Colorectal Surgery Service | 594 | 575 | 614 | 5.1 | 4.7 | 5.5 |
| Hepatobiliary and Pancreatic Surgery Service | 1,309 | 1,253 | 1,365 | 14.6 | 13.3 | 15.9 |
| Upper Gastrointestinal Surgery Service | 678 | 653 | 703 | 4.1 | 3.5 | 4.7 |
| Vascular Surgery Service | 453 | 423 | 482 | 7.4 | 6.7 | 8.1 |
| Spinal Surgery Service | 818 | 786 | 851 | 15.1 | 14.3 | 15.9 |
| Trauma and Orthopaedic Service | 307 | 298 | 317 | 6.7 | 6.5 | 6.9 |
| Ear Nose and Throat Service | 328 | 314 | 343 | 2.3 | 2.0 | 2.6 |
| Ophthalmology Service | 143 | 122 | 164 | 5.1 | 4.6 | 5.6 |
| Oral Surgery Service | -4 | -18 | 10 | -1.5 | -1.8 | -1.1 |
| Neurosurgical Service | 956 | 922 | 990 | 23.1 | 22.2 | 23.9 |
| Plastic Surgery Service | 144 | 124 | 164 | 1.7 | 1.3 | 2.1 |
| Cardiac Surgery Service | 1,728 | 1,619 | 1,838 | 24.5 | 22.0 | 27.0 |
| Thoracic Surgery Service | 1,071 | 990 | 1,153 | 14.1 | 12.0 | 16.2 |
| Anaesthetic Service | 475 | 391 | 559 | 18.8 | 16.5 | 21.0 |
| Pain Management Service | 257 | 237 | 278 | 12.6 | 12.0 | 13.2 |
| General Internal Medicine Service | 524 | 500 | 547 | 5.0 | 4.5 | 5.5 |
| Gastroenterology Service | 229 | 218 | 240 | -0.3 | -0.6 | -0.1 |
| Endocrinology Service | 1,668 | 1,611 | 1,725 | 3.6 | 2.4 | 4.9 |
| Clinical Haematology Service | 1,805 | 1,754 | 1,857 | 17.1 | 16.0 | 18.2 |
| Hepatology Service | 464 | 374 | 554 | 8.9 | 6.8 | 11.0 |
| Cardiology Service | 579 | 553 | 606 | 8.1 | 7.5 | 8.6 |
| Dermatology Service | 354 | 326 | 383 | 1.9 | 1.3 | 2.4 |
| Respiratory Medicine Service | 382 | 341 | 423 | 7.9 | 7.0 | 8.8 |
| Renal Medicine Service | 896 | 809 | 983 | 14.8 | 12.9 | 16.6 |
| Medical Oncology Service | 1,239 | 1,180 | 1,299 | 18.2 | 16.9 | 19.5 |
| Neurology Service | 535 | 495 | 574 | 11.0 | 10.0 | 12.0 |
| Rheumatology Service | 271 | 228 | 314 | 6.5 | 5.3 | 7.8 |
| Clinical Oncology Service | 1,680 | 1,631 | 1,730 | 29.9 | 28.7 | 31.1 |
| Interventional Radiology Service | 444 | 406 | 483 | 8.5 | 7.6 | 9.3 |

**Supplementary Table 2. Cumulative treatments effects on earnings and employment across all 32 treatment specialities**

| **Treatment speciality** | **Cumulative treatment effect (£)** | **Cumulative treatment effect (person-years)** |
| --- | --- | --- |
| General Surgery Service | 8,753,552,144 | 215,031 |
| Urology Service | 5,523,304,056 | 19,538 |
| Breast Surgery Service | 1,527,487,254 | 13,298 |
| Colorectal Surgery Service | 6,232,633,902 | 41,437 |
| Hepatobiliary and Pancreatic Surgery Service | 1,033,643,754 | 9,269 |
| Upper Gastrointestinal Surgery Service | 2,480,587,050 | 12,971 |
| Vascular Surgery Service | 1,021,992,480 | 13,430 |
| Spinal Surgery Service | 1,617,848,517 | 23,214 |
| Trauma and Orthopaedic Service | 6,883,785,690 | 123,635 |
| Ear Nose and Throat Service | 2,812,264,771 | 17,641 |
| Ophthalmology Service | 1,022,177,250 | 29,328 |
| Oral Surgery Service | 13,774,362 | - 7,419 |
| Neurosurgical Service | 1,747,269,225 | 33,439 |
| Plastic Surgery Service | 846,093,768 | 8,398 |
| Cardiac Surgery Service | 875,037,198 | 7,476 |
| Thoracic Surgery Service | 507,054,142 | 4,331 |
| Anaesthetic Service | 121,243,878 | 4,582 |
| Pain Management Service | 1,050,789,568 | 42,802 |
| General Internal Medicine Service | 2,729,770,812 | 20,318 |
| Gastroenterology Service | 7,541,943,900 | - 50,470 |
| Endocrinology Service | 1,624,506,306 | 2,130 |
| Clinical Haematology Service | 4,153,678,730 | 19,178 |
| Hepatology Service | 176,674,103 | 2,622 |
| Cardiology Service | 3,210,987,347 | 32,167 |
| Dermatology Service | 1,358,758,950 | 5,990 |
| Respiratory Medicine Service | 726,248,190 | 11,569 |
| Renal Medicine Service | 486,509,130 | 5,674 |
| Medical Oncology Service | 2,151,121,432 | 20,202 |
| Neurology Service | 691,741,336 | 9,846 |
| Rheumatology Service | 306,049,352 | 4,901 |
| Clinical Oncology Service | 3,620,616,040 | 36,307 |
| Interventional Radiology Service | 899,992,224 | 12,428 |

**Supplementary appendix: Further method details**

Patients get older during the study period, and age is a key determinant of pay and employment status. Therefore, age was a confounder of the relationship between the exposure variable of interest (time before/since treatment) and the outcome variable of interest (pay/employment) and had to be controlled for in the analysis. This was done by standardising monthly values of mean pay and probability of employment, such that the age distribution (measured in half-year intervals) in each month was held fixed at that in Month 0; in other words, we derived the expected pay/employment in each month if patients remained at the same age as when they were treated (or in the case of the Census sample, at the randomly assigned index date).

After applying the initial inclusion criterion of patients being aged 25-64 years at treatment, the dataset was further filtered to include only monthly data when patients were aged ≥30 years and <59 years when computing the age-standardised values. This was to ensure that people in all half-year age bands that were present in the study population in Month 0 had the opportunity to contribute to the age-standardised estimates in all months before and after Month 0, before "ageing out" of the age range covered for follow-up. For example, if the upper age limit in Month 0 was 64 years, the maximum observed age in the study cohort would be 59 years by Month -60. Similarly, if the lower age limit in Month 0 was 25 years, the minimum observed age in the study cohort would be 30 years by Month 60. Therefore, an increasing number of half-year age bands would become empty with time backwards or forwards from Month 0, despite these age bands carrying a weight >0 in the age-standardisation process in these months (as they are populated in Month 0), hence the age-standardised estimates would appear artificially low. By restricting the analytical dataset to person-months when patients are aged 30 to <59 years, we guaranteed that the half-year age groups that comprised the standard population (i.e. those that are present in Month 0) also had the opportunity to be represented in every month over the 10-year period. This is demonstrated in Figure 1.1 below.

Figure 1.1: Demonstration of the need to further restrict the initial study cohort (comprising individuals aged 25-64 years at treatment) based on age at each month of follow-up

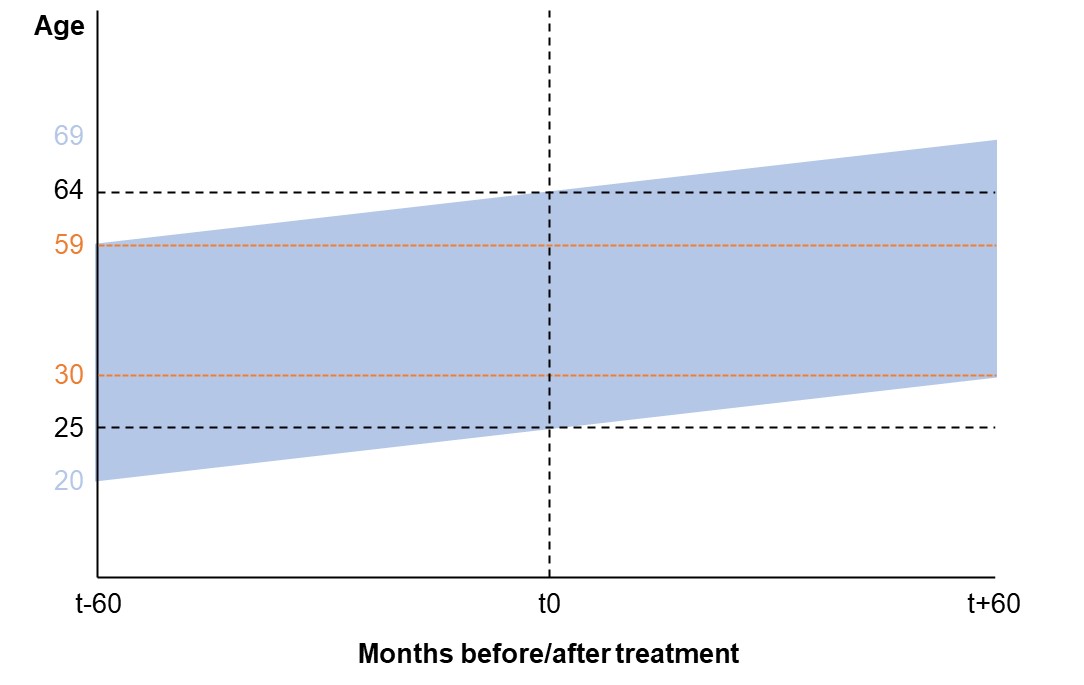

For each treatment specialty in each inpatient/outpatient setting, monthly values of mean pay and probability of employment were calculated in 58 half-year age bands from [≥30.0 years to <30.5 years] to [≥58.5 years to <59.0 years]. Age-standardised values of pay/employment in each month were then calculated as a weighted average of the 58 age-specific values, where the weights were proportional to the size of the study population in each half-year age band in Month 0:

$y_{j,k,t}^{*}=\sum_{a=1}^{58} y_{a,j,k,t}s_{a,j,k}$	[1a]

$s_{a,j,k}={n_{a,j,k,0}}/{\sum_{a=1}^{58} n_{a,j,k,0}}$	[1b]

where $y_{j,k,t}^{*}$ is the age-standardised value of pay/employment for treatment specialty $j$ in inpatient/outpatient setting $k$ in Month $t$; $y$ is mean pay or percentage of patients in paid employment; $s_{a}$ is the weight (population share in age band $a$) to use in the weighted average; and $n_{0}$ is the cell population size in Month 0.

When comparing age-standardised values between a particular treatment specialty and the general population Census sample, it should be remembered that the cohorts are standardised to two different standard populations (i.e. the age distribution of each of the two respective cohorts in Month 0). Therefore, although the age profile will be held constant through time for each of the cohorts, it will not be the same between the cohorts, and differences in the levels of age-standardised pay/employment between the cohorts could be partly driven by differences in age structure.

In order to capture uncertainty inherent to both the age-standardised estimates and the counterfactual values of pay/employment, confidence intervals were generated via simulation (it was not possible to perform bootstrapping from the observed study data for computational reasons). Within each of the 24 study populations, for each of the 121 time periods (Month -60 to Month 60), 10,000 values of pay among paid employees were simulated from a log-normal distribution with ‘location’ and ‘scale’ parameters determined from the observed study data:

${Pay}_{j,k,t} \sim Lognormal({Location}_{j,k,t}, {Scale}_{j,k,t})$	[2a]

${Location}_{j,k,t}=log\left[ \frac{m_{j,k,t}^{2}}{\sqrt{s_{j,k,t}^{2}+m_{j,k,t}^{2}}} \right]$	[2b]

${Scale}_{j,k,t}=\sqrt{log\left[ 1+(s_{j,k,t}^{2}/m_{j,k,t}^{2}) \right]}$	[2c]

where $m$ and $s$ are the sample mean and standard deviation of pay among paid employees, respectively.

For each time period, we also simulated 10,000 probabilities of being a paid employee from a binomial distribution, again with ‘number of trials’ and ‘probability of success’ parameters determined from the observed data:

${Employment}_{j,k,t} \sim Binomial(n_{j,k,t}, {\sum_{i=1}^{n_{j,k,t}} E_{i,j,k,t}}/{n_{j,k,t}})$	[3]

where $E_{i}$ = 1 if patient $i$ is a paid employee and 0 otherwise.

We then multiplied the simulated values for pay among paid employees by the simulated probabilities of being a paid employee to derive simulated values of pay among all working-age people (including those who were not paid employees).

For each time period, the 10,000 simulated values of pay and probability of being a paid employee were age-standardised, the standard error [$\hat{SE}(y^{*})$] was estimated as the standard deviation of the simulated values, and 95% confidence intervals were constructed as:

$95\% CI: y_{j,k,t}^{*}\pm1.96\times\hat{SE}(y_{j,k,t}^{*})$	[4]

Model fitting

In order to derive estimates of treatment and exposure effects, linear regression models were fitted to age-standardised values of pay/employment over a span of the pre-treatment period. Although we computed age-standardised trajectories back to Month -60, we only used the one-year period before treatment, i.e. Month -12 to Month -1, for model fitting. As the study period for HES inclusion began on March 2015 and the PAYE data went back as far as March 2014, the period Month -12 to Month -1 was the only pre-treatment period for which all included study patients had the opportunity to contribute to the estimated trajectory. For example, patients whose index date was in March 2015 could not have contributed to the estimate for Month -13, as PAYE records were not available for February 2014. Trends in age-standardised values prior to Month -12 may therefore have partly reflected changes in the composition of the cohort (e.g. if there was heterogeneity in the trajectory by calendar time of treatment), as well as genuine disease-related changes in pay/employment.

The one-year period before treatment was conceptually divided into two epochs: ‘pre-disease’ (i.e. not yet ill) and ‘pre-treatment’ (i.e. ill but not yet treated). It was impossible to directly identify disease onset from the HES data alone, so this had to be inferred from the data by locating a breakpoint in the age-standardised pay/employment trajectory (i.e. the point at which illness started to impact on individuals’ ability to work). If a breakpoint was located at Month $x$ (-11 ≤ $x$ ≤ -2), the period from Month -12 to Month [$x$-1] was inferred to be the ‘pre-disease’ epoch, and the period from Month $x$ to Month -1 was inferred to be the ‘pre-treatment’ epoch.

The location of the breakpoint was automatically identified using R’s ‘segmented’ package^3^. This package uses an empirical search strategy to find the value of $breakpoint$ that minimises $\left| \hat{\beta}_{3} \right|$ in the segmented regression model:

$\hat{y}_{j,k,t}^{*}=\hat{\beta}_{0,j,k}+\hat{\beta}_{1,j,k,t}+\hat{\beta}_{2,j,k,t}{(t-{breakpoint}_{j,k})}_{+}+\hat{\beta}_{3,j,k,t}\left( {gap}_{j,k} \right)$	[5]

where ${(t-{breakpoint}_{j,k})}_{+}=(t-{breakpoint}_{j,k})$ if $t>{breakpoint}_{j,k}$ and 0 otherwise; and ${gap}_{j,k}=1$ if $t>{breakpoint}_{j,k}$ and 0 otherwise. The estimated coefficient $\hat{\beta}_{1}$ represents the slope of the pre-breakpoint trajectory, $\hat{\beta}_{2}$ represents the change in slope after the breakpoint, and $\hat{\beta}_{3}$ represents the size of the discontinuity between the pre- and-post breakpoint trajectories. Separate models were estimated, and therefore separate breakpoints were identified, for pay and employment.

In the simplified example below (Figure 3.2), there are three candidate breakpoints: Month -3, Month -7 and Month -10 (in reality, the ‘segmented’ package considered all values along the continuous number line from Month -2 to Month -11 as candidate breakpoints). Month -7 would be chosen as the optimised location of the breakpoint, as this is the specification that minimises the size of the discontinuity between the pre- and post-breakpoint segmented.

Figure 1.2: Illustrative example of how the ‘segmented’ R package chooses the optimal location of a breakpoint by minimising the gap between pre- and post-breakpoint segments

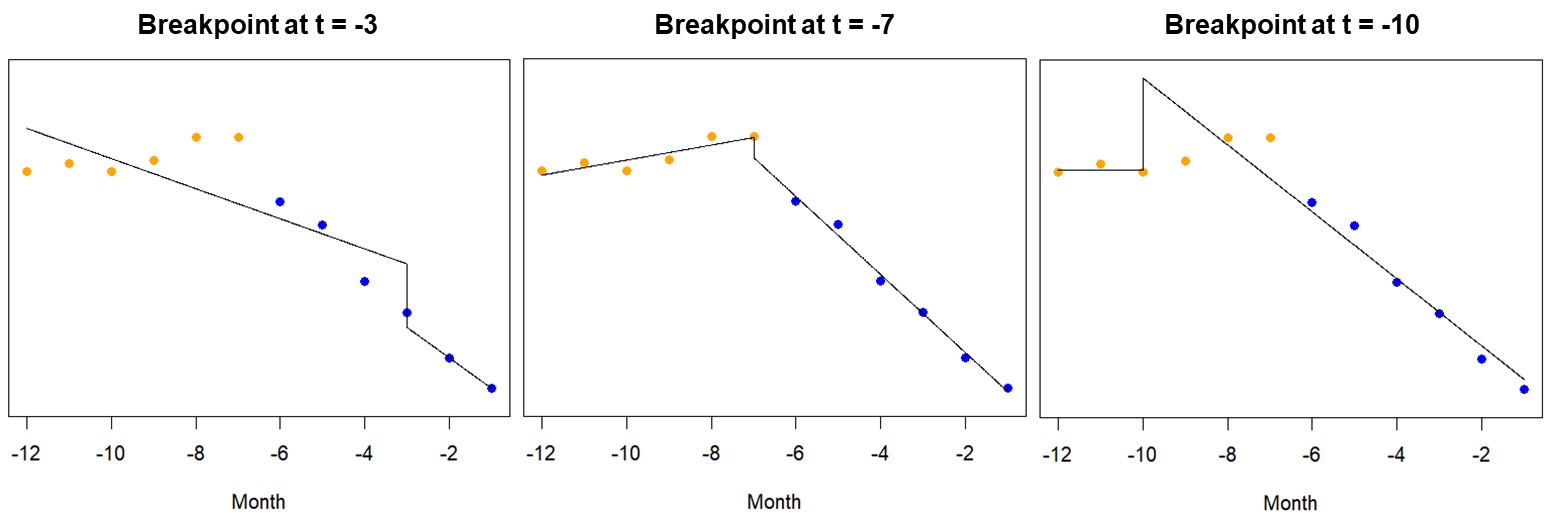

Once the breakpoint was identified, the $gap$ variable was removed from the model described in equation [5] so that the fitted values did not exhibit a discontinuity. Therefore, the final model was:

$\hat{y}_{j,k,t}^{*}=\hat{\beta}_{0,j,k}+\hat{\beta}_{1,j,k,t}+\hat{\beta}_{2,j,k,t}{(t-{breakpoint}_{j,k})}_{+}$	[6]

In addition to performing a breakpoint analysis with linear regression models, we also explored second- and third-order polynomial regression models fitted to the entire one-year pre-treatment span, thus allowing for a gradual rather than an abrupt change in the trajectory. Although these models sometimes provided a better fit to the training data than the linear breakpoint models, they often resulted in “explosive behaviour” in their out-of-sample projections, with estimated values of mean pay and probability of paid employment rapidly tending to zero after Month -1. These models were therefore not considered further.

Treatment and exposure effects

The treatment effect was estimated as the difference between the observed and model-projected values of age-standardised pay/employment in each month following treatment:

$\Delta_{j,k,t}=y_{j,k,t}^{*}-\hat{y}_{j,k,t}^{*}$ for $0\leq t\leq60$	[7]

The projected values represent a counterfactual outcome; that is, the expected trajectory of age-standardised pay/employment had individuals not received treatment and instead remained in their ‘pre-treatment’ state. This interrupted time series (ITS) design^4^ is illustrated in Figure 3.3 below.

Figure 1.3: Stylised illustration of how treatment effects are estimated

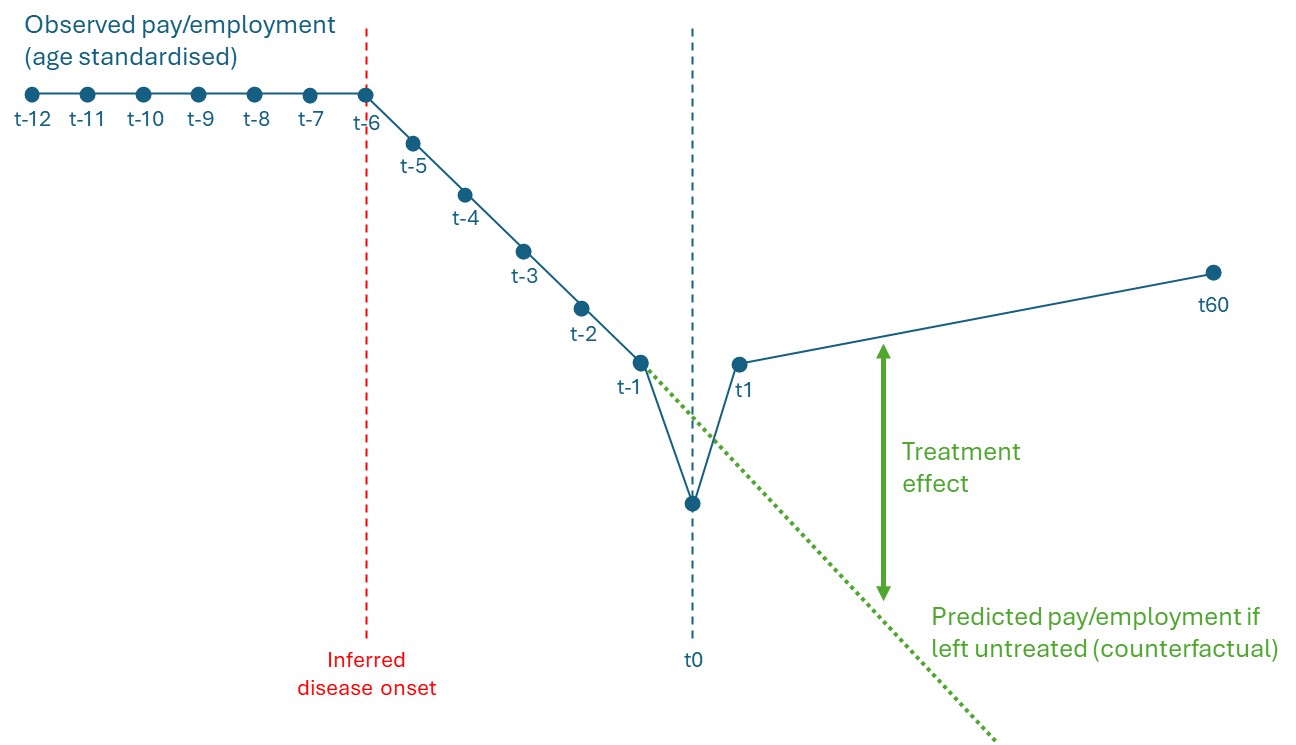

For each of the 24 study populations, confidence intervals were produced around the estimated treatment effects by fitting the model described in equation [6] to each of the 10,000 simulated, age-standardised pay/employment trajectories (as described earlier); calculating the corresponding 10,000 sets of treatment effects; estimating the standard error ([$\hat{SE}(\Delta)$]) for each month as the standard deviation of the 10,000 treatment effects; and constructing the 95% confidence intervals as:

$95\% CI: \Delta_{j,k,t}\pm1.96\times\hat{SE}(\Delta_{j,k,t})$	[8]
